# Advancing cardiovascular disease risk prediction beyond conventional methods: a systematic review of multimodal machine learning models integrating traditional clinical factors and multi-omics data

**DOI:** 10.1101/2025.10.07.25337473

**Authors:** Lyns Etienne, Pierre Bauvin, Alaedine Benani, Maryne Lepoittevin, Sylvain Bodard

**Affiliations:** Department of Chemical Biology, Harvard University, Cambridge MA 02138; Cancer Program, Broad Institute of MIT and Harvard, Cambridge MA 02142; Preventive Medicine, Data Science and AI Lab, Zoī, F-75010, Paris, France; Service de médecine vasculaire, hôpital européen Georges Pompidou (HEGP), AP-HP, Université Paris-Cité, Paris, France; Sorbonne Université, Université Sorbonne Paris Nord, INSERM, Limics, 75006 Paris, France; Memorial Sloan Kettering Cancer Center (MSK), Department of Radiology, Cornell University, 1275 York Avenue, New York, NY 10065, USA; Transplantation Sciences, Massachusetts General Hospital, Harvard Medical School, Boston, USA; Université de Paris Cité, Hôpital Universitaire Necker Enfants Malades, Service d’Imagerie Adulte, AP-HP, F-75015, Paris, France; CNRS UMR 7371, INSERM U 1146, Laboratoire d’Imagerie Biomédicale (LIB), Sorbonne Université, F-75006, Paris, France

## Abstract

**Background:** Cardiovascular disease (CVD) is a leading global health burden. Traditional risk prediction models, though widely used, often overlook genetic predisposition and other complex biological factors, which significantly impacts CVD risk. The emergence of multi-omics technologies now enables a more comprehensive view of an individual’s risk, but integrating such high-dimensional data has been challenging and requires advanced computational approaches. Recent advances in machine learning methods now offer powerful tools to synthesize and integrate these high-throughput dataset, offering a promising approach to improve CVD risk stratification.

**Objective:** This systematic review assesses whether CVD risk prediction models incorporating omics data alongside clinical and other variables improve prediction compared to using clinical or omics data alone.

**Methods:** A systematic search was conducted across PubMed (MEDLINE), Embase, and Web of Science databases in June 2025 using keywords related to CVD, risk prediction, multi-omics data, and machine learning. Studies reporting on models comparing multi-omics data with traditional clinical factors for CVD risk prediction were included. Data on model performance, methodologies, and subgroup analyses were extracted and synthesized.

**Results:** Studies consistently showed that clinical models integrating multiple modalities, including approximately genomic (n=58), biomarkers (n=109), biological (n=125), and other data types significantly enhanced CVD risk prediction, with combined clinical+genomic models outperforming single-modality approaches. Other data types like lifestyle factors and proteomics further refined performance. Subgroup analyses revealed decreased predictor accuracy across diverse ancestries and age-specific performance differences. Importantly, genetically defined high-risk individuals often derived greater absolute benefits from targeted clinical interventions. Models effectively spanned from predicting risk in asymptomatic individuals for primary prevention to guiding prognosis in diseased patients for secondary prevention

**Conclusion:** CVD risk prediction models integrating genomic, clinical, and other variables offer superior accuracy and refined stratification. These advanced models hold immense potential for personalized interventions across diverse populations. Future research should prioritize real-world implementation and broad validation to translate these findings into routine clinical practice.

## 1. Introduction

Cardiovascular disease (CVD) remains the leading cause of morbidity and mortality worldwide, accounting for an estimated 640 million people living with some form of CVD and 20.5 million CVD-related deaths in 2025 alone.^1–3^ CVD encompasses disorders of the heart and blood vessels, with common types including coronary artery disease (CAD), heart failure (HF), atherosclerotic cardiovascular disease (ASCVD), stroke, peripheral artery disease (PAD), and arrhythmias such as atrial fibrillation (AF). These conditions affect individuals of all ages, with a higher prevalence among adults aged 40 and older, particularly men. Even though younger adults have a lower incidence of CVDs, modifiable risk factors like obesity, hypertension, and diabetes contribute more significantly to CVD development in younger than in older adults.^4^ This distinction is critical, as traditional age-based risk prediction models may underestimate risk in younger populations, highlighting the need for earlier identification and intervention to reduce long-term CVD burden. The CVD global burden is expected to worsen over the next two decades, reaching a projected 30.5 million deaths annually.^4^ This underscores a critical need for effective strategies to identify individuals with moderate to elevated risk before the onset of disease.^2,4^ The underlying causes of CVD are complex and multifactorial, involving a combination of genetic predispositions, environmental exposures, lifestyle behaviors, demographic characteristics, and other health and risk factors.^1^ Early CVD diagnosis through risk prediction models can enable timely intervention and targeted prevention, which are essential for reducing the incidence and impact of CVD.

Traditionally, CVD risk prediction has relied on clinical models that incorporate established risk factors such as age, sex, blood pressure, body mass index (BMI), cholesterol levels, smoking status, and diabetes.^5^ Widely used risk assessment tools such as the Framingham Risk Score^6^ and the ASCVD Risk Estimator^7^ have informed prevention guidelines and clinical decision-making for years. However, these models do not currently account for genetic predisposition, which has been shown to contribute significantly to individual variation in cardiovascular risk.

Advancements in next-generation sequencing (NGS) technologies have allowed scientists to generate large amounts of multi-omics data, including genomics, transcriptomics, proteomics, metabolomics, which led to the discovery of many genetic variants associated with CVD.^8^ The use of NGS technologies has led to the development of polygenic risk scores (PRS), which are composite measures of inherited risk derived from the cumulative effect of numerous genetic variants from single nucleotide polymorphisms (SNPs) across the genome. Several PRS have been developed for different types of CVD, which have been reviewed previously.^9^ The CVD PRS have already been shown to provide additional insights to the pre-disposition of certain groups of people to various types of CVD, notably offering the potential to improve CVD risk stratification by identifying individuals who may be at high genetic risk despite having few or no traditional risk factors.^10^ To effectively analyze and integrate this high-dimensional, complex data, machine learning algorithms have emerged as powerful tools. These advanced computational approaches can identify intricate patterns and non-linear relationships within vast datasets, potentially yielding more accurate and nuanced risk predictions than traditional statistical methods.

Recently, a growing number of studies have explored the integration of multi-omics data with traditional clinical risk factors, lifestyle information, and environmental variables in machine learning-based predictive models to assess CVD risk.^11^ The rationale is that combining multi-omics, clinical, and lifestyle information in advanced machine learning models may yield more accurate and personalized estimates of CVD risk in both healthy individuals and diseased patients, particularly in early or subclinical stages. Additionally, genomic information could help identify patients that are at high risk and are more likely to develop cardiovascular events and could benefit from lifestyle interventions and medical treatment. The objective of this systematic review is to evaluate whether risk prediction models that incorporate genomic data alongside clinical and other variables (e.g., lifestyle, environments) improve the prediction of future cardiovascular disease occurrence compared to models using clinical variables or genomic data alone.

## 2. Methods

### 2.1. Protocol and Registration

This systematic review was conducted in accordance with the Preferred Reporting Items for Systematic Reviews and Meta-Analyses (PRISMA) guidelines (**Supplementary Method 1**) to ensure methodological rigor and transparency.^12^ A study protocol was created and uploaded to the PROSPERO database (**Supplementary Method 2**). The review protocol was prospectively registered in the PROSPERO database of systematic reviews (registration number: CRD420251086792).

### 2.2. Data Sources and Literature Search Strategy

A comprehensive literature search was conducted on June 3, 2025, across three electronic databases: PubMed (MEDLINE), Embase, and Web of Science, to identify studies that developed or validated CVD risk prediction models incorporating genomic data and applying machine learning, artificial intelligence or related analytical techniques. The search strategy combined terms related to cardiovascular conditions, genomics, risk prediction, model performance (e.g., AUC, ROC), and artificial intelligence or machine learning.

The search was restricted to English-language publications. In total, the search yielded 643 records, which were deduplicated to 504 unique articles for screening. The detailed literature search strategy for each database is provided in (**Supplementary Table 1**).

### 2.3. Review Methods and Study Selection

We considered only studies that were original research. The screening was completed in two stages. Title and abstract screening was conducted using Rayyann^13^, a web-based tool for systematic review screening. One reviewer (LCE) screened all articles at the title and abstract level. After this initial screening, 73 studies met the inclusion criteria and were retained for full-text review. When full-text articles could not be retrieved through institutional access or public databases, corresponding authors were contacted directly to obtain copies of the manuscripts. Two reviewers (LCE and ML) conducted the second stage of review and 40 studies met the eligibility criteria. The screening results were exported from Rayyan as a CSV file for further documentation and analysis in Microsoft Excel. (**Supplementary Table 2**) All included studies focused on the development or validation of CVD risk prediction models. Common features among these studies included the integration of diverse data types (such as clinical, genomic variables), an emphasis on predictive accuracy, and the use of standardized performance metrics. Most studies also incorporated internal and/or external validation techniques to assess the robustness of their models. Studies were grouped by cardiovascular disease type and summarized in structured tables presenting key model features, performance, and validation details.

### 2.4 Eligibility Criteria

Studies were eligible for inclusion if they developed or validated multivariable prediction models for CVD risk in adult populations. Models had to incorporate genomic, or other omics data, such as PRS, DNA methylation, transcriptomic profiles, proteomic or metabolomic data, either alone or in combination with clinical, lifestyle, or environmental variables. Eligible study designs included cohort studies, case-control studies, cross-sectional studies, or model validation studies. To be included, studies were required to report at least one model performance metric, such as area under the curve (AUC), concordance index (C-index), calibration score (e.g., Brier score), net reclassification index (NRI), or integrated discrimination improvement (IDI). The target population included adults either at risk for CVD or previously diagnosed with CVD. Studies using prediction models based solely on clinical data were not excluded if they were used as comparators to omics-based models. Only peer-reviewed articles published in English within the defined time frame were considered. Studies were excluded if they focused solely on diagnosis, prognosis, or if they were reviews, meta-analyses, editorials, commentaries, or conference abstracts.

### 2.5 Data Collection and Extraction

Data were extracted from the 40 included studies that met the eligibility criteria. Key characteristics extracted included the type of cardiovascular condition, data sources used (e.g., clinical exam, electronic health records, demographic, medical imaging, biological or biochemical, genomics), model features, performance metrics, and validation approaches.

Studies were grouped by cardiovascular disease type and summarized in structured tables presenting core information on model inputs, methodology, performance, and validation. Most studies reported performance using standardized metrics and included either internal or external validation to benchmark against existing prediction models. All code generated to produce the figures are provided in the supplementary materials (**Supplementary Method 3**).

Data were extracted using a standardized form developed for this review. For each included study, the following information was recorded and are discussed in the later sections (**Supplementary Tables 3 and 4**):

- Study characteristics: title, authors, publication year, and study design.
- Population details: eligibility criteria, CVD subtype studied, dataset origin, and sample size.
- Model characteristics: type of prediction model (e.g., logistic regression, machine learning, deep learning), variables included (clinical, genomic, transcriptomic, epigenomic, metabolomic, proteomic), and whether the model was newly developed, externally validated, or both.
- Outcome and Performance metrics: primary outcome (e.g., incident CVD, myocardial infarction, stroke), AUC, C-statistic, calibration (e.g., Brier score), NRI, sensitivity, specificity, and other relevant metrics.
- Validation type: internal validation (e.g., cross-validation, bootstrapping) or external validation using independent datasets or cohorts.
- Risk of bias and transparency: availability of the model or code, efforts to address overfitting, handling of missing data, and clarity and completeness of model reporting.

During the preparation of this work, we used several AI tools to improve the efficiency and consistency of our data extraction and synthesis. NotebookLM (Google) was used for initial data extraction and organization, summarizing key information from the source documents. SciSpace was employed to identify and extract relevant sections from PDFs, such as model descriptions, validation details, and results tables. DocAnalyzer.ai (GPT-4.1-mini, OpenAI) was used to assist with automated document parsing, enabling rapid comparison of methodological elements and performance metrics across studies. Finally, ChatGPT (version GPT-4o-2024-08-06 and GPT-4o1, OpenAI) was used to refine the manuscript’s wording, clarify extracted features, and improve overall readability. All outputs generated by these tools were meticulously reviewed and verified by the study authors to ensure accuracy and appropriateness, with final approval given before inclusion in the study.

### 2.6. Risk of Bias Assessment

Studies will be assessed using a simplified version of the Prediction model Risk Of Bias ASsessment Tool (PROBAST)^14^ tool across four domains: participants, predictors, outcomes, and analysis. Each domain will be rated as low, high, or unclear risk of bias.

## 3. Results

### 3.1 Study Selection

The study selection process is summarized in Figure 1 (PRISMA flowchart).

**Figure 1.** Systematic review PRISMA flow diagram of study inclusions and exclusions. The chart presents the number of records identified, screened, assessed for eligibility, and included in the review, with corresponding reasons for exclusion at each stage

### 3.2 Study Characteristics

The characteristics of included articles, such as study populations, model features, performance and outcome measures are reported in later sections. (**Supplementary Tables 3 and 4**).

### 3.3 Overview of predictive models by cardiovascular disease type

Among the 40 studies included in this systematic review, the most commonly represented cardiovascular condition was CAD, featured in 17 studies (42.5%). Stroke-related outcomes (including ischemic stroke and stroke as a CVD risk factor) were examined in 7 studies (17.5%), followed by AF in 6 studies (15%). ASCVD and myocardial infarction (MI), including ST-segment elevation myocardial infarction (STEMI) and non-ST-segment elevation myocardial infarction (NSTEMI), were each represented in 5 studies (12.5%). HF and composite outcomes such as major adverse cardiovascular events (MACE) and major adverse cardiac and cerebrovascular events (MACCE) appeared in 3 studies each (7.5%). Other less frequently reported conditions included hypertension, hypertrophic cardiomyopathy (HCM), ischemic heart disease (IHD), and PAD (n = 2, 5% each). Single studies reported on intracerebral hemorrhage (ICH), extracranial artery stenosis (ECAS), critical limb ischemia (CLI), Long QT Syndrome (LQTS), and generalized atherosclerosis. Two studies did not specify a cardiovascular disease subtype. A summary of the CVD subtypes represented across the studies is included in Table 1.

**Table 1.** Summary of the various CVD subtypes across the different studies reviewed.

| Cardiovascular Disease (CVD) Type | N | % |
| --- | --- | --- |
| Coronary artery disease (CAD) | 17 | 42.5 |
| Stroke | 7 | 17.5 |
| Atrial fibrillation (AF) | 6 | 15 |
| Atherosclerotic cardiovascular disease (ASCVD) | 5 | 12.5 |
| Myocardial infarction (MI/STEMI/NSTEMI) | 5 | 12.5 |
| Heart failure (HF) | 3 | 7.5 |
| Major adverse cardiovascular events (MACE/MACCE) | 3 | 7.5 |
| Hypertrophic cardiomyopathy (HCM) | 2 | 5 |
| No subtype specified (generic CVD) | 2 | 5 |
| Hypertension (CVD risk factor) | 2 | 5 |
| Peripheral artery disease (PAD/CLI) | 2 | 5 |
| Ischemic heart disease (IHD) | 2 | 5 |
| Intracerebral hemorrhage (ICH) | 1 | 2.5 |
| Long QT Syndrome (LQTS) | 1 | 2.5 |
| Critical Limb Ischemia (CLI) | 1 | 2.5 |
| Extracranial artery stenosis (ECAS) | 1 | 2.5 |
| Atherosclerosis (unspecified) | 1 | 2.5 |

### 3.4 Genomic data incorporation

The studies reviewed primarily incorporated genomic data using SNPs and PRS, largely derived from comprehensive genome-wide association studies (GWAS) summary statistics. Genomic data were integrated through various methods, including machine learning techniques for selecting predictive SNP subsets and constructing classifiers for disease risk prediction. For instance, the Young Finns Study employed a machine-learning-based SNP-subset selection procedure to identify informative genetic variants for carotid atherosclerosis risk prediction.^15^ Similarly, several studies leveraging the UK Biobank database developed genetic predictors using SNP genotype data to predict both biomarkers and disease risk. Widen *et al.* developed PGS predictors for 65 blood and urine markers, with these scores demonstrating strong correlations with certain traits. The predictor for lipoprotein A (LpA) levels, a highly heritable risk factor for heart disease, achieved a correlation of approximately 0.76. The study also developed biomarker risk scores (BMRS) to predict common disease risk from biomarkers alone. For conditions like CAD risk, a BMRS achieved an AUC of approximately 0.75, and for diabetes, it reached an AUC of about 0.95.^16^

PRS derivation commonly involves additive models based on large GWAS summary statistics. Castela Forte *et al.* computed PRSs for CAD, type 2 diabetes (T2D), and hypertension using GWAS results constructed from the UK Biobank data. They applied linkage disequilibrium (LD) correction tools like LDpred to adjust SNP weights and minimize inflation caused by correlated SNPs.^17^ In a study on East Asian CAD patients, PRS were optimized for predicting all-cause death using GWAS summary statistics for 15 traits from Biobank Japan.^18^ Chen *et al.* used PRS combined with phenome-wide association study (PheWAS)-derived risk factors to build predictive models, leveraging tools like PLINK and Bayesian regression methods.^19^ Some meta-prediction frameworks explicitly excluded UK Biobank-derived PRS metrics to prevent overfitting, instead utilizing large-scale GWAS data to develop generalizable genetic models with superior accuracy and calibration across diverse populations.^20^

Ancestry-specific considerations were addressed in some studies, though the need for further diversification and validation across various ancestries was often noted. The meta-prediction model by Chen *et al.* was validated across European and African American populations.^20^ UK Biobank-based studies primarily focused on European ancestry groups, employing imputation quality control measures to ensure the reliability of genotype data.^17^ The East Asian CAD study inherently reflected an ancestry-specific genetic architecture by using GWAS summary statistics from Biobank Japan.^18^ Although some studies integrated genetic risk scores with conventional risk factors to improve prediction across ancestries, detailed, explicit adjustments for ancestry beyond cohort selection and validation were limited.^16,20^

### 3.5 Comparative performance of prediction models

Direct comparisons of models based solely on clinical or biological variables, those utilizing only omics data, and integrated models combining both clinical or biological, and genomic information consistently showed that including genomic or biomarker data, particularly SNPs or PRSs, or proteomic and microbiome data enhanced CVD risk prediction.

Castela Forte *et al.* showed that incorporating genetic susceptibility scores, physical measurements, and other biomarkers into decision rules models improved risk prediction for CAD, T2D, and hypertension when compared to clinical models like the Framingham Risk Score (FRS).^17^ For CAD, the decision rule model outperformed the FRS in men (AUC 0.66 vs 0.60) but not in women. For T2D, AUC improved to 0.75 vs 0.72, and for hypertension to 0.70 vs 0.60. High-risk groups identified by the decision rule model had markedly elevated disease incidence (40-fold for CAD, 40.9-fold for T2D, 21.6-fold for hypertension), with significant reclassification gains (NRI: 5.8% for T2D, 19.9% for hypertension).

Omiye *et al.* showed that adding PAD-specific PRS to clinical models improved prediction performance for PAD and consistently outperformed clinical-only models across various metrics, including AUC, NRI, IDI, and decision curve analysis (DCA).^21^ Specifically, for PAD detection, AUC increased from 0.902 (clinical-only) to 0.909 (clinical+PRS), with improved calibration (Brier score 0.172 to 0.169). Reclassification was significant (categorical-NRI 0.07, 95% CI 0.002–0.137; p=0.04), especially in the intermediate-risk group, and discrimination improved (IDI 0.0092, p<0.001). Qin *et al.* observed significant improvement in predicting all-cause death in East Asian CAD patients when a metaPRS model was combined with clinical risk factors.^18^

The authors evaluated a metaPRS (integrating CAD and 8 risk-factor PRSs) in East Asian CAD patients. The clinical model alone (age, sex, smoking) had an AUC of 0.72. The metaPRS alone achieved AUC 0.63, and the combined clinical+metaPRS model improved performance to AUC 0.76, with +4% sensitivity at the same specificity versus clinical-only. Risk stratification showed clear gradients: high-risk patients (≥3rd quartile) had a 3.99-fold increased risk of death (95% CI 2.40–6.64), and intermediate-risk patients a 2.18-fold increased risk (95% CI 1.33–3.59), compared with the low-risk group. Chen *et al.* built a meta-prediction model integrating multiple PRSs and clinical and biometric data for CAD risk prediction, achieving an area under the receiver operating curve (AUROC) of 0.84, and a 67% higher area under the precision-recall curve (AUPRC), indicating enhanced risk stratification a notable improvement over other methods (the 0.72–0.79 AUROC).^20^ Møller *et al.* reported that adding a CAD-specific PRS (PRSCAD) to a clinical prediction model, PROMISE minimal risk score (PMRS) increased the AUC from 0.76 to 0.79 for predicting the absence of CAD, leading to better reclassification in low-to-intermediate risk groups.^22^ Mazidi *et al.* observed modest but measurable enhancements in C-statistics and NRIs for ischemic heart disease prediction in both European and Chinese populations when PRS were added to conventional risk factors.^23^ In the Chinese cohort, the C-statistic was 0.845 with conventional factors and 0.846 with PS added (minimal change). In the European cohort, C-statistics improved from 0.725 to 0.734, with a significant NRI of 46.7% (95% CI 38.9–54.6).

Mishra *et al.* found that a coronary artery calcification genetic risk score (CAC GRS) offered additional predictive value and improved CAD prediction accuracy when combined with a broader CAD metaGRS.^24^ Specifically, the authors evaluated a CAC GRS (11 SNPs) alongside a broader CAD metaGRS (1.7M variants) in the LURIC cohort (n=2742). The clinical risk factor model had an AUC of 0.717. Adding the CAC GRS improved AUC to 0.734 (p=0.02), while adding the metaGRS raised it to 0.784 (p=0.001). Combining both scores with clinical factors yielded AUC 0.787. Widen *et al.* emphasized the benefits of integrating genomic and biomarker data for improved disease risk prediction, including CAD.^16^ The BMRS achieved good discrimination, with AUC ∼0.75 for CAD and ∼0.95 for diabetes. An ASCVD BMRS using ∼10 biomarkers performed as well as or better than the ACC ASCVD Risk Estimator. Climente-González *et al.* indicated that while proteomics-based models initially outperformed clinical scores and PRSs alone, combining proteomics with clinical features further improved prediction.^25^ The proteomics-only model achieved AUROC 0.767, AUPRC 0.241, compared with PREVENT/QRISK3 (0.736), SCORE2 (0.674), PCE (0.666), and PRSs (0.547–0.577).

Combining proteomics with clinical features further improved performance (AUROC 0.785, AUPRC 0.284), a +2.0% AUROC and +9.3% AUPRC gain over a clinical-only model (0.770, 0.259). Syed *et al.* demonstrated that integrating a CHD PRS with an AI-derived retinal image risk score (predMACE10) significantly enhanced the prediction of MACE beyond clinical risk scores alone.^26^ The PCE and retinal score alone each had AUC 0.697, while the PRS alone performed worse (AUC 0.575). A model combining retinal score, PRS, and clinical factors (age, sex, HbA1c, diabetes duration) outperformed PCE (AUC 0.686 vs. 0.658; p<0.001). The most comprehensive model (retinal + PRS + PCE) achieved the highest discrimination (AUC 0.728, 95% CI 0.695–0.761).

Niu *et al.* developed a PRS (PGGRS) from 13 SNPs and evaluated it in a rural Chinese cohort.^27^ Models with traditional risk factors alone achieved AUCs of 0.785 (Cox), 0.790 (ANN), 0.838 (RF), and 0.854 (GBM). Adding the PGGRS improved performance, raising AUCs to 0.861 (RF) and 0.871 (GBM), confirming improved discrimination with the PGGRS addition.

Reclassification gains were substantial, with NRI 44.9% (RF) and 22.9% (GBM), and IDI ∼4.7% for both. Han *et al.* constructed a novel 69-variant PRS (HF-PRS) for heart failure with preserved ejection fraction (HFpEF), that outperformed conventional scores (MAGGIC and ASCEND-HF) for 1-year mortality prediction with an AUC of 0.852, further improving to 0.880 when combined with clinical variables.^28^ For AF, Chen *et al.* and Jabbour *et al.* integrated PRSs with phenomic, clinical, and EHR data, demonstrating improved early detection and risk stratification.^19,29^ Chen *et al.* showed that a PRS alone achieved an AUC of 0.600, which increased to 0.855 (P<0.001) when combined with age and sex, and PheWAS analyses confirmed top associations with cardiovascular diseases. Similarly, Jabbour *et al.* found that an AF-PRS alone had an AUC of 0.59, compared with 0.62 for the CHARGE-AF clinical score and 0.76 for an ECG-AI model; integrating the PRS and CHARGE-AF with ECG-AI modestly improved the AUC to 0.77, and risk stratification showed high-risk groups with 4.3-fold (ECG-AI) and 1.85-fold (PRS) increased incidence of AF. In hypertrophic cardiomyopathy (HCM), SVEP1, a novel biomarker, was integrated with NT-proBNP and clinical features to predict MACE, increasing the C-statistic from 0.82 to 0.87.^30^ Similarly, Liu *et al.* showed that combining gut microbiome markers with clinical variables improved STEMI prediction in young males (AUC = 0.877; validated AUC = 0.934).^31^ A distribution of the AUC performance across all the studies reviewed is shown in Fig. 2. These studies collectively reinforce the conclusion that multi-omics integration, whether genomic, proteomic, transcriptomic, or microbiomic, enhances risk stratification and supports more precise, personalized prediction models across a spectrum of CVD conditions.

**Figure 2.**
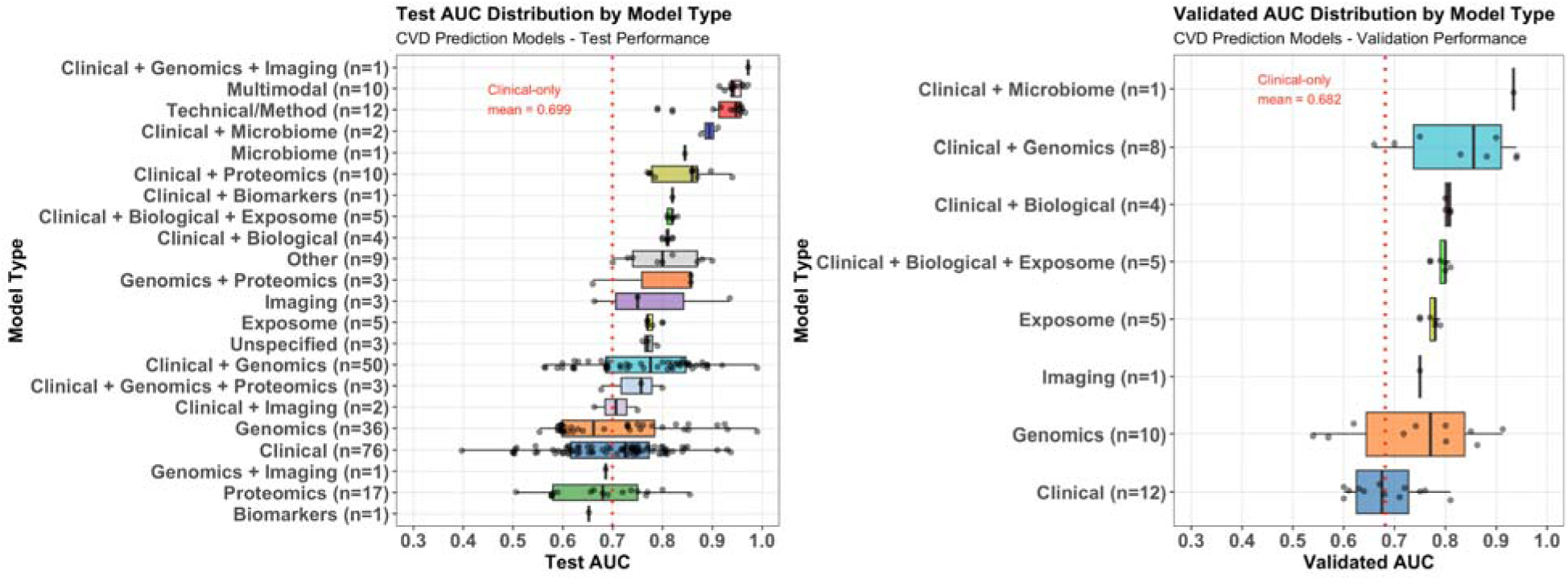
Distribution of AUC performance by model type in cardiovascular disease prediction studies. (A) Left panel shows AUC values for models with both internal and external validation. (B) Right panel presents models with external validation only. Individual AUC values are displayed as dots, with boxes indicating the spread of values for each model type. The red dotted lines is a visual guide showing the average performance of clinical-only models as a reference.

While most evidence supports the added benefit of genomic or biomarker data into CVD risk prediction models, the degree of improvement can vary (Fig. 2), and some studies have reported no significant improvement or inconsistent results. Møller *et al.* observed that individual proteins in the PMRS did not significantly enhance predictive performance for the absence of CAD.^22^ Biener *et al.* reported that while some microRNAs (miRNAs) were associated with acute coronary syndrome (ACS), they did not provide added diagnostic value for NSTEMI over established biomarkers like high-sensitivity cardiac troponin T (hs-cTnT).^32^ Oguz *et al.* reported improved predictive accuracy when combining clinical variables with SNP genotypes in one cohort, but this improvement did not replicate in an independent cohort, emphasizing the challenge of generalizability and highlighting potential inconsistencies across different populations.^33^ The performance of PRS can decrease across different ancestries, highlighting the need for diverse datasets and validation in varied populations.^34^ Christensen *et al.* evaluated SNP-based and combined models for AF, venous thromboembolism (VTE), and pneumonia.

SNP-only models performed poorly (AUC 0.60 for VTE, 0.60 for AF, and ∼0.50 for pneumonia). Adding SNPs to clinical models yielded only modest and inconsistent gains: for AF, AUC improved from 78.7% to 80.0% (linear regression) and 78.8% to 79.8% (deep learning); for VTE, from 63.4% to 66.6% (linear), but no improvement with deep learning; for pneumonia, AUC slightly decreased from 69.2% to 68.4% (linear regression); 69.7% (unchanged for deep learning). Overall, genetics contributed limited incremental predictive value across phenotypes in these models.^35^ Together, these findings underscore that while integration of genomic and biomarker data can enhance CVD risk prediction, the benefits are context-dependent and must be interpreted with consideration for population characteristics, model design, and the interplay with traditional risk factors.

### 3.6 Subgroup stratification

Several studies explicitly categorize patients into distinct cardiovascular risk levels, often leveraging advanced multi-modal models. Mordi *et al.* stratified individuals with T2D using a retinal risk score derived from retinal vascular parameters, which independently predicted MACE (HR per SD increase=1.11, 95% CI 1.04–1.18). When stratified by tertiles, patients in the highest tertile of the retinal risk score had a significantly increased risk of MACE compared to those in the lowest tertile (HR 1.32, 95% CI 1.13–1.55, P < 0.001). The retinal score with age and sex achieved an AUC of 0.663, comparable to the Pooled Cohort Equations (PCE) (0.658), while combining the retinal score with a CHD PRS significantly improved discrimination (AUC=0.686, P<0.001) and reclassification (NRI=0.240, IDI=0.019, both P<0.001), with modest further gains when the PCE was added (AUC=0.690). This combined model also yielded significant reclassification improvements (NRI=0.240, IDI=0.019, both P<0.001).^36^ Castela Forte *et al.* developed decision rule models that classified individuals into high-risk tiers for CAD, diabetes, and hypertension, demonstrating substantially higher hazard ratios (e.g., 40-fold for CAD) compared to traditional clinical risk calculators.^17^ Chen *et al.* presented a meta-prediction model for CAD that significantly improved risk stratification across all percentiles of predicted risk over a 10-year follow-up, which was notably higher than existing clinical and polygenic risk scores.^20^ In the UK Biobank test set, the model achieved an AUROC of 0.84 (95% CI 0.83– 0.85) and an AUPRC of 0.38, compared with 0.73 for PCE, 0.75 for QRISK3, and 0.72 for PREVENT. The model stratified risk effectively, with CAD incidence ranging from 0.3% in the lowest risk percentile to 62.9% in the highest. It also improved reclassification, with NRIs of 0.20 over PCE, 0.14 over QRISK3, and 0.21 over PREVENT. External validation in the All of Us cohort confirmed strong performance (AUROC = 0.81 vs. 0.72–0.73 for conventional scores).

Møller *et al.* highlighted the incremental value of integrating genomic and proteomic data with clinical factors for CAD risk classification.^22^ In 1,462 patients with chest pain, clinical risk factors alone predicted high-risk coronary plaques with an AUC of 73.2, while a multi-PGS alone achieved 60.1 and proteomics alone 69.0. Combining clinical factors with the polygenic score improved prediction to an AUC of 74.8 (P = 0.004), whereas adding proteomics provided no further benefit. These studies collectively demonstrate that combining clinical scores (e.g., PCE, QRISK, Framingham) with PRS, imaging, biomarkers, and proteomic data enables robust patient classification into cardiovascular risk tiers, showing strong associations with biological markers and clinical outcomes.

A few studies explicitly assessed whether individuals at different levels of genomic or biomarker-defined risk derive differential benefit from interventions. For example, Chen *et al.* demonstrated that individuals with high polygenic risk for CAD achieved significantly greater absolute risk reductions from standard interventions than low-genetic-risk individuals. For LDL cholesterol lowering to 35 mg/dL, high-risk individuals had a 7.10% absolute risk reduction versus 0.39% in low-risk individuals, and at 100 mg/dL, 4.34% versus 0.33%. For HbA1c lowering to 5.6%, high-risk individuals achieved a 4.89% reduction versus 2.02% in low-risk individuals, and at 6%, 3.90% versus 1.37%. For systolic blood pressure targets of 110 mmHg, the absolute risk reduction was 2.71% versus 1.94% in low-risk individuals, and at 120 mmHg, 2.58% versus 1.84%. These results underscore the clinical utility of genetically-informed treatment targets for maximizing cardiovascular risk reduction.^20^

As previously mentioned, Castela Forte *et al.* showed that a decision rules model integrating polygenic scores and biomarkers could better stratify individuals for lifestyle interventions in preventive care, such as weight loss, limiting saturated fat, and 30 minutes of daily exercise, aimed at reducing CVD risk and progression to type 2 diabetes. Their model identified high-risk individuals who were more likely to develop CAD, hypertension, or diabetes and reduced the number of unnecessary interventions compared to traditional risk scores such as the FRS, while still capturing most incident cases.^17^ Specifically for clinical utility, the model would recommend lifestyle interventions to 41.6% fewer individuals for T2D and 53% fewer for hypertension than the FRS, with NRI of 5.8% and 19.9%, respectively. The HCM study by Osawa *et al.* highlighted that plasma SVEP1 levels provided added risk stratification beyond NT-proBNP, identifying high-risk subgroups who might benefit from earlier or more aggressive management (e.g., anticoagulation or weight loss).^30^ In 610 HCM patients, high SVEP1 predicted increased risk of MACE even with low NT-proBNP (HR 4.52, 95% CI 1.05–19.4), while high levels of both biomarkers had the highest risk (HR 10.4, 95% CI 2.81–38.4). Adding SVEP1 to clinical predictors and NT-proBNP improved the C-statistic from 0.82 to 0.87, and a model with SVEP1 alone outperformed NT-proBNP (C-statistic 0.86 vs. 0.82), highlighting its utility for identifying high-risk patients. This supports a precision prevention framework where clinical decisions, including interventions, are tailored to molecularly defined risk profiles. While many other studies explored risk prediction, they did not directly examine how stratification affects intervention outcomes, pointing to a gap in the current literature.

Several studies compared subgroups based on population characteristics like ancestry and age, revealing differences in predictive performance and clinical outcomes. Chen *et al.* found that their meta-prediction model, which integrated PRS and meta-features, was more effective than age and sex alone in stratifying CAD risk subgroups. The meta-prediction model achieved an AUC of 0.84 (95% CI 0.83–0.85). In comparison, a model based solely on age and sex had a much lower AUC of 0.75 The study also noted that genetic risk significantly influenced the effectiveness of clinical intervention across these subgroups.^20^ For individuals with a high genetic risk of CAD, lowering LDL cholesterol to a target of 35 mg/dL resulted in an absolute risk reduction of 7.10%, compared to just 0.39% for those with low genetic risk. For patients with an HbA1c ≥6%, those with a high genetic risk for type 2 diabetes achieved a 2.53% greater absolute risk reduction from HbA1c lowering compared to those with low genetic risk (3.90% vs 1.37%). For individuals with a systolic blood pressure (SBP) ≥140 mm Hg, those with a high SBP PRS had a 0.74% greater absolute risk reduction (2.58% vs 1.84%) from lowering SBP to a target of 120 mm Hg compared to those with a low SBP PRS. Widen *et al.* observed that polygenic predictor accuracy decreased with increasing genetic distance between training and testing ancestries, highlighting the impact of population substructure.^16^ While the predictors were trained exclusively on data from individuals of European ancestry, they were evaluated across different self-reported ancestry groups. For example, the performance (correlation) of the PGS for Lipoprotein A (LpA), which was the best-performing predictor, showed the following pattern across different ancestries: European: ∼0.76, South Asian: ∼0.35, East Asian: ∼0.12, and African: ∼0.10. Møller *et al.* found improved prediction accuracy (AUC 66.5%) in younger patients (≤55 years old) compared to those >55 years old (AUC 58.3%) for high-risk coronary plaques using non-protein models and GPSMult.^22^ Kany *et al.* developed the Predict-AF3 model integrating clinical (CHARGE-AF), genetic (PRS), and ECG-AI data to predict 5-year AF risk.

Predict-AF3 achieved the highest discrimination (AUC 0.817, 95% CI 0.802–0.832) compared with CHARGE-AF+ECG-AI (AUC 0.802), CHARGE-AF alone (0.785), ECG-AI alone (0.705), and PRS alone (0.618). Using the Predict-AF3 model, individuals were stratified into risk categories.^37^ : cumulative AF incidence was 5.83% in the high-risk group (>2.5% predicted), 1.51% in the intermediate group (1-2.5%), and 0.56% in the low-risk group (≤1%). When all three models classified individuals as high risk, incidence reached 11.06%, compared with 0.67% when none did.

The predictive models vary in their target populations, assessing CVD risk in both asymptomatic healthy individuals and patients with existing CVD or related conditions. Many models, often utilizing large population cohorts like the UK Biobank, prioritize primary prevention, aiming to identify risk in individuals without a prior CVD diagnosis through methods such as proteomics-based Explainable Boosting Machine (EBM) models and integrated genetic and clinical factors.^20,25,38^ Conversely, other models concentrate on secondary prevention or prognosis for those already diseased, as seen in models classifying CVD in nonalcoholic fatty liver disease (NAFLD) patients or predicting MACE in high-risk individuals with T2D.^26,39–41^

### 3.7 Clinical features, input variables, and performance metrics

Clinical variables used across studies consistently included traditional risk factors: demographic (age, sex)^36,39,41^, anthropometric measures (weight and body mass index (BMI))^39^, blood pressure^36,41^, comprehensive lipid profile (total cholesterol, low-density lipoprotein cholesterol (LDL-c), high-density lipoprotein cholesterol (HDL-c), and triglycerides)^24,39,41^, glucose metabolism markers (fasting glucose and hemoglobin A1c (HbA1c)).^39,41^ Lifestyle factors (smoking status, alcohol consumption, and physical activity) were frequently incorporated.^38,39^ Some studies also include inflammatory markers (high-sensitivity C-reactive protein (hs-CRP)) and medication use.^39^ Other laboratory parameters such as liver enzymes, creatinine, hemoglobin, platelet count, and urate levels are included in specific cohorts, especially when investigating populations with specific comorbidities like NAFLD.^39^

Various studies identified specific variants, proteins, and clinical factors as key predictors. For AF, 30 important SNPs were isolated from, primarily on chromosomes 10 and 16, involving genes such as NEURL1, SH3PXD2A, INA, NT5C2, STN1, and ZFHX3. Notably, INA, NT5C2, and STN1 were newly linked to AF.^19^ In ICH, circERBB2, circCHST12, and hsa_circ_0005505 were identified as promising biomarkers.^42^ For ischemic stroke functional outcome, rs11109607 (ANKS1B) was found to be the most significant genetic variant.^38^ In CAD prediction, key exposome attributes were identified using SHapley Additive exPlanations (SHAP).^38^ Li et al. found that circulating plasma hsa-miR-4739 was significantly elevated in T2DM patients with critical limb ischemia and independently associated with disease risk (adjusted OR 12.8, 95% CI 1.15–143.1, P=0.038). ROC analysis showed modest diagnostic performance for hsa-miR-4739 alone (AUC 0.69, sensitivity 61.5%, specificity 73.3%), but combining it with clinical risk factors substantially improved discrimination (AUC 0.94 vs. 0.91 for risk factors alone).^43^

Beyond genomics (PRS, SNPs), other data types positively influenced model performance. Integrating clinical (e.g.; waist circumference), lifestyle (e.g., sedentary behavior) and specific SNPs (IL16 and ANKLE1 genes), along with hypertension, in an ensemble-based random forest model for CVD classification in NAFLD patients from the UK Biobank cohort achieved an AUC of 0.849 (95% CI, 0.840–0.855). This integrative model outperformed models with fewer data types, such as those using only clinical data (AUC 0.799), only lifestyle data (AUC 0.652), or only genetic data (AUC 0.617).^39^ Incorporating genome-wide genetic liabilities, derived from hundreds of thousands of genetic variants, modestly improved stroke risk classification.^44^ PRS and SNPs data consistently improved model performance for CAD and other cardiovascular outcomes when compared to traditional risk scores.^16,33^ For example, a machine learning approach to predict hypertension using SNPs, age, and gender achieved an AUC of 89% (88.5% for the neural network), compared with ∼81% for age+gender alone and only 50–51% for gender alone, showing a clear gain in accuracy from adding genomic data.^45^ Similarly, another study found that adding a PRS for CAD to a clinical risk model improved the AUC for predicting CAD by 3%.^34^ Proteomic data significantly improved heart failure prediction accuracy.^46^ A prediction model for HFpEF using PRS (HF-PRS) of 69 genetic variants achieved an AUC of 0.852 (95% CI 0.827–0.877), which was a 15.6% improvement over a clinical risk score model that had an AUC of 0.696 (95% CI 0.658–0.734). The addition of the HF-PRS resulted in a NRI of 74.1% (p < 0.001) and an IDI of 18.1% (p < 0.001).

Various ML algorithms, including linear regression, neural networks (NNs), random forests (RF), decision trees (DT), support vector machines (SVM), K-nearest neighbor (KNN), logistic regression (LR), Gaussian Naive Bayes (GNB), XGBoost, and LightGBM, along with ensemble methods, have been employed.^27,33,39,42,45^ Model evaluation across the studies employed various performance metrics, with AUC being the most common for assessing model discrimination.

Examples include studies by Smole *et al.* which reported an AUC of 0.82 for their machine learning-based risk stratification model for hypertrophic cardiomyopathy (HCM-RSS).^47^ A novel attention-based cross-modal transfer learning framework for CVD prediction achieved an AUC of 0.972.^48^ Widen *et al.* noted AUC values ranging from 0.81 to 0.82 for various machine learning models predicting CVD risk.^16^ Chen *et al.* reported an AUROC of 0.84 for a meta-prediction model of CAD risk.^20^ Sharma *et al.* found an AUC of 0.849 for CVD prediction NAFLD in patients.^39^ Climente-González *et al.* reported AUROC values of 0.767 for proteomics-only models and 0.785 for combined models with clinical features for CVD risk prediction.^25^ For survival analysis, the C-index was a primary metric, with Chen *et al.* reporting a C-index of 0.82 for a survival-based XGBoost estimator.^20^ Calibration metrics like the Brier Score and ICI were also used, as shown in studies by Chen et al.^20^ and MacCarthy and Pazoki^44^, respectively.

Other metrics included the F1 Score, combining precision and recall (0.71) by Smole et al.^47^ Sensitivity and specificity were reported by Chen *et al.* as 76.6% and 76.2%, respectively.^20^ The NRI and IDI were utilized to quantify the improvement in risk prediction achieved by adding genomic markers or proteomics data.^21,25^ Precision and recall were also reported in a study by Prakash *et al.*, 92.0% and 94.5% respectively.^48^

While AUC is a widely used metric that allows for some level of comparison across studies, direct comparability is often limited by inherent differences in cohort characteristics, outcome definitions, follow-up periods, and the types of models employed. Nevertheless, the use of additional metrics such as the C-index for survival models, calibration measures (Brier score, calibration plots), and reclassification indices (NRI, IDI) provides a more comprehensive evaluation of predictive performance. Studies like Chen *et al.* specifically highlighted improved calibration and discrimination when compared to existing clinical risk scores, suggesting a benchmarking against established models.^20^ The frequent use of large datasets, particularly the UK Biobank, across several studies introduces some consistency in data sources, but variations in disease focus (e.g., HCM, AF, CAD, stroke) and modeling approaches still influence the direct comparability of reported metrics.^16,20,21,25,29,39,44^

### 3.8 Risk of bias

The risk of bias assessment revealed a varied but predominantly high risk of bias across the included studies. (Fig. 3) While the Predictors domain showed a consistently low risk of bias in all studies (100%), the Participants, Outcome, and Analysis domains all had a considerable portion of studies with a high risk of bias. Overall, 28 studies (70%) were classified as high risk, with the Analysis domain identified as the most significant source of bias at 82.5% (33 studies). Given the pervasive nature of these biases, the findings of this systematic review should be interpreted with caution.

**Figure 3.**
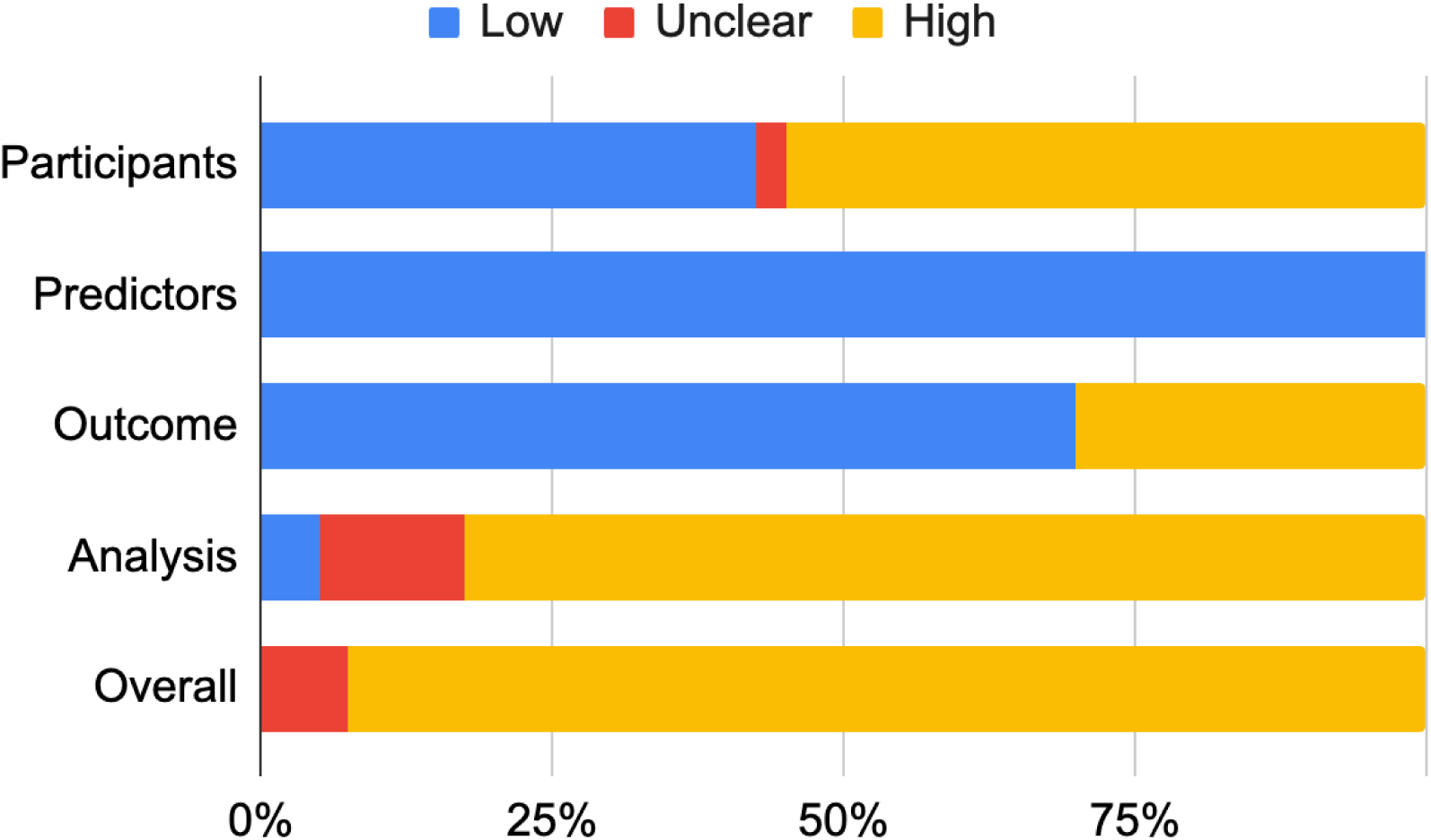
Risk of bias assessment. Risk of bias for included studies, categorized by domain: Participants, Predictors, Outcome, Analysis, and Overall. The bars represent the percentage of studies with a low (blue), unclear (red), or high (yellow) risk of bias. The overall risk of bias is a summary of the individual domain assessments.

## Discussion

The integration of diverse data types and advanced ML techniques holds significant potential for clinical utility. The reviewed literature consistently demonstrates the growing integration and value of genomic data, primarily through the use of SNPs and PRS, in refining CVD risk prediction. The development of these genetic scores, often based on large-scale genomic studies, and enhanced by ML approaches, allows for a more personalized assessment of an individual’s predisposition to various cardiovascular conditions. This approach, including the creation of generalizable meta-prediction models, highlights the progress in maximizing genetic utility. The general consensus from these studies is that combining genomic data with traditional clinical risk factors significantly enhances the accuracy of CVD risk prediction models. This “hybrid” approach often outperforms models relying solely on either clinical or genomic information, demonstrating a synergy where each data type contributes unique and valuable insights.^17,18,20,21^ The ability of these integrated models to more accurately identify individuals across the entire CVD risk spectrum, from very high to very low, holds substantial promise for tailoring preventive strategies and interventions for CVD patients. The practical benefits of incorporating genomic information are further evidenced by demonstrable improvements in key performance metrics such as AUC and reclassification indices.^20,22–24,26^ These advancements could empower clinicians to identify at-risk individuals earlier and with greater precision, leading to more timely and effective interventions.

Beyond genomic data, the strategic incorporation of other data types like lifestyle factors, and proteomics further strengthens predictive models.^22,23,25,46^ These additions, complementing traditional clinical variables, contribute to a comprehensive, multi-modal approach to CVD risk assessment. This holistic strategy allows for a more nuanced understanding of underlying CVD mechanisms, providing richer insights than any single data type could offer alone. Continuous research into these diverse data sources and their optimal integration through advanced analytical techniques promises to refine our ability to predict, prevent, and manage CVD more effectively. Improved risk stratification allows for personalized interventions for AF and the identification of high-risk HFpEF patients who may benefit from intensive management strategies.^19,28^ Decision rule models, when integrated with digital data or digital therapeutics platforms, can support the successful adoption of lifestyle interventions in preventive and primary care for common chronic conditions like CAD, T2D, and hypertension.^17^ Early detection and prevention of CVD are enhanced through the use of blood protein markers.^25^ An AI approach allows for rapid retinal CVD risk assessment in routine clinical care, potentially reducing the need for multiple healthcare attendances and saving time on regular blood pressure and cholesterol monitoring.^26^

Despite these advancements, several limitations and areas for future research remain. It’s crucial to acknowledge the ongoing need for careful consideration of ancestry-specific differences, which, while addressed in some studies through cohort selection and external validation, warrant more detailed explicit adjustments to ensure equitable performance across all populations.^17,18,20,23^ While the degree of improvement varies with the specific condition and population, the consistent trend towards enhanced predictive power strongly indicates the value of integrating genomic insights into clinical practice. It is important to acknowledge that not every genomic or biomarker addition will yield significant improvements in all contexts. The few instances where combined models offered limited or inconsistent advantages serve as a valuable reminder that rigorous validation and a nuanced understanding of a marker’s incremental value within a specific clinical setting are essential.^22,32,33^ Moreover, the studies highlight the critical importance of considering population characteristics, including ancestry and age, in both the development and application of cardiovascular risk prediction models. The observed decrease in predictive accuracy across different ancestries underscores the necessity for models trained and validated on diverse populations to ensure equitable performance.^16^

Similarly, age-specific performance differences suggest that the utility of risk factors and models may vary across age ranges, potentially requiring age-tailored approaches.^22^ Crucially, evidence indicates that genetic risk can influence the differential benefit individuals derive from interventions, reinforcing the value of precise subgroup stratifications for personalized care.^20^ It is crucial to acknowledge that predictive features do not necessarily imply a causal relationship, thus necessitating further validation through experimental approaches and independent cohorts to confirm causality. Additionally, the use of PRS or genetics data in risk prediction models has several limitations as previously discussed, and those must be taken into account, during both research and clinical application.

Finally, the predictive models reviewed target diverse populations, ranging from asymptomatic healthy individuals for primary prevention to patients with existing CVD or related conditions for secondary prevention and prognosis. This distinction in target populations is critical for the clinical utility of these predictive models. Models developed for asymptomatic populations hold immense potential for early intervention and primary prevention, identifying individuals who might benefit most from lifestyle modifications or targeted therapies before disease onset.

Traditional clinical risk scores, often serving as benchmarks, primarily align with this primary prevention objective. Conversely, the inclusion of already diseased patients in some models is equally valuable, as these tools can help refine disease management, predict adverse events, and guide prognosis in high-risk or established patient cohorts. Some models even bridge the gap between primary and secondary prevention by identifying high-risk individuals, such as those with T2D, who may appear asymptomatic but are predisposed to developing MACE, as demonstrated in recent deep learning approaches.^26^ Ultimately, the diverse scope of these models, ranging from general population screening to specialized patient management, underscores the breadth of applications for advanced predictive analytics in cardiovascular health, each serving a distinct yet complementary role in improving patient outcomes.

The evolving landscape of cardiovascular disease risk prediction is ultimately moving towards a more comprehensive and personalized approach. These advanced data modalities often provide complementary information not captured by traditional clinical risk factors alone. While individual genomic models might not always outperform comprehensive clinical models alone, their integration often leads to statistically significant enhancements in performance metrics like AUC, NRI, and IDI, allowing for more precise risk stratification and identification of high-risk individuals for targeted interventions The enduring challenge lies in translating these sophisticated research findings into accessible and actionable tools for routine clinical care, thereby ensuring that the benefits of precision medicine are broadly realized.

## Supporting information

PRISMA

## Data Availability

All data produced in the present study are available upon reasonable request to the authors

## Supplementary Information

- Supplementary Table 1 (Literature Search Strategy): <u>Supplementary Materials_FINAL</u>
- Supplementary Table 2 (Screening Results): <u>Supplementary Materials_FINAL</u>
- Supplementary Table 3 (Data Extraction): <u>Supplementary Materials_FINAL</u>
- Supplementary Table 4 (Models Characteristics): <u>Supplementary Materials_FINAL</u>
- Supplementary Method 1 (PRISMA guidelines): <u>Supplementary Materials_FINAL</u>
- Supplementary Method 2 (Prospero Study Protocol): <u>Supplementary Materials_FINAL</u>
- Supplementary Method 3 (Analysis Code): <u>Supplementary Materials_FINAL</u>
- Risk of bias tool/template (e.g., PROBAST)

## Acknowledgments

The authors are grateful to Zoī for its support and resources that contributed to the successful completion of this systematic review. Its commitment to advancing research and innovation has been instrumental. The authors would like to thank Paul Bain for helpful discussion and providing feedback regarding the literature search.

## Registration

NIHR-PROSPERO CRD420251086792

## Ethics approval statement

This study is a systematic review. It did not involve any human subjects, patient data, or new data collection, and therefore did not require approval from an Institutional Review Board or Ethics Committee.

